# Genetically determined body mass index mediates the effect of smoking on type 2 diabetes risk, but not coronary artery disease risk

**DOI:** 10.1101/2020.01.30.20019737

**Authors:** Christopher S Thom, Zhuoran Ding, Michael G Levin, Scott M Damrauer, Benjamin F Voight

## Abstract

Clinical observations have linked tobacco smoking with increased type 2 diabetes risk (1–5), a major public health concern (6). Mendelian randomization analysis has recently suggested smoking may be a causal risk factor for type 2 diabetes (7). However, this initial association could be mediated by additional causal risk factors correlated with smoking behavior, which have not been investigated to date. We hypothesized that body mass index (BMI) could explain the association between smoking and diabetes risk. First, we confirmed previous reports that genetically determined smoking behavior increased risk for both type 2 diabetes (OR=1.21, 95% CI: 1.15-1.27, P=1×10^−12^) and coronary artery disease (CAD; OR=1.21, 95% CI: 1.16-1.26, P=2×10^−20^). Additionally, a 2-fold increased smoking risk is positively associated with body mass index (BMI; ∼0.8 kg/m^2^, 95% CI: 0.54-0.98 kg/m^2^, P=1.8×10^−11^). In multivariable Mendelian randomization analysis, including BMI accounted for nearly all of the risk of smoking on type 2 diabetes (OR 1.06, 95% CI: 1.01-1.11, P=0.03). In contrast, the independent association between smoking and CAD persisted (OR 1.12, CI: 1.08-1.17, P=3×10^−8^) despite controlling for BMI. Causal mediation analyses agreed with these estimates. Our findings support a model whereby smoking initiation increases obesity, which in turn increases type 2 diabetes risk, with minimal if any direct effects from smoking on diabetes risk. Patients should be advised to stop smoking to limit both type 2 diabetes and CAD risk, and therapeutic efforts should consider pathophysiology relating smoking and obesity.

## Introduction

Obesity and type 2 diabetes are leading causes of death worldwide (6). These conditions and related comorbidities are increasing global epidemics, expected to place ever larger demands on health care systems. As such, revealing the underlying causal risk factors and their associated biological pathways are crucial for prevention to public health.

Epidemiologic (1–5) studies have associated smoking with increased diabetes risk. Indeed, smoking can perturb glycemic regulation (4,8). However, smoking also impacts a number of cardiometabolic traits, including obesity, blood pressure and heart disease (9–11). Some studies have shown that BMI increases smoking risk (9), while others have suggested that smoking may actually decrease BMI (10,12). An improved understanding of the mechanisms underlying the link between smoking and diabetes may inform targeted therapeutic development and clinical decision making.

Recent collections of genetic data for smoking behavior, type 2 diabetes, and cardiometabolic factors such as body mass index (BMI) provide vehicles through which evidence of causality can be evaluated, using the framework of Mendelian randomization (MR). Formally, MR uses genetic variants associated with an exposure of interest (i.e., smoking behavior) to create an instrumental variable to estimate a potential causal effect of genetically-determined exposure on an outcome (i.e., type 2 diabetes). Because alleles are randomly allocated at meiosis, and due to the fact that genotype precedes phenotype, this approach with care can address issues of confounding and reverse causality that limit inference in prospective or cross-sectional cohort studies (13). Multivariable MR and causal mediation analyses extend the method to identify and account for additional causal factors that may mediate univariable experimental analyses (14,15).

Here, we used MR analysis on summary-level statistics from individuals of European ancestry to define relationships between smoking and cardiometabolic traits. We hypothesized that the relationship between smoking and type 2 diabetes was mediated by BMI, and in this study aimed to (i) estimate the effect of genetically predicted smoking traits on type 2 diabetes, (ii) compare the effect of smoking behavior on type 2 diabetes with the effect of smoking on coronary artery disease (CAD), (iii) estimate the effect of smoking on BMI, and (iv) determine if BMI mediates the effects of smoking on type 2 diabetes and/or CAD. Our findings elucidate causal genetic effects between these clinically important traits. Better understanding these biological associations will inform clinical management and aid translational research efforts.

## Results

### Increased genetically determined odds of smoking initiation elevates type 2 diabetes risk

First, we assessed if genetically determined smoking initiation, smoking cessation, and smoking heaviness (number of cigarettes per day) modulated susceptibility to type 2 diabetes. To estimate a causal effect between odds of smoking initiation and susceptibility to type 2 diabetes, we performed causal inference analysis using MR. Using summary statistics obtained from genome-wide association studies of these traits, we generated an instrumental variable (IV) for smoking initiation (16), and tested its association with type 2 diabetes (17). Our genetic instrument comprised 341 linkage-independent single nucleotide polymorphisms (SNPs, EUR r^2^<0.01) that met genome-wide significance for this smoking behavior. This instrument was not subject to weak instrument bias, nor were other instrumental variables used in this study (**Supplemental Table 1**) (18). We observed that both inverse variance weighted (P=1.0×10^−12^) and median weighted (P=6.9×10^−12^) MR methods demonstrated statistically significant positive association between increased genetically determined odds of smoking initiation and susceptibility to type 2 diabetes (**Fig. 1A**). As a sensitivity analysis, we performed the MR-Egger regression test, and found no evidence of systematic bias in our estimated effect (MR-Egger intercept term P=0.88). These data support the hypothesis that increased odds of smoking initiation corresponds with an increase of type 2 diabetes; each 2-fold increase in genetic predisposition to smoking initiation corresponding to a 21% increased odds of type 2 diabetes risk (OR=1.21, 95% confidence interval = 1.15-1.27, by inverse variance weighted method). This effect size was consistent with a recent study (7).

**Table 1.** Genetic correlations between smoking traits (smoking initiation, Smk Init; smoking cessation, Smk Cess; smoking heaviness, Cig-per-day; lifetime smoking, LfSmk), body mass index (BMI), type 2 diabetes (T2D), HbA1c, and coronary artery disease (CAD) based on LD Score Regression (20).

| Traits | | Genetic Correlation ($r_g$) | P-Value |
| --- | --- | --- | --- |
| Between smoking traits |  |  |  |
| Smk Init | Smk Cess | 0.391 | $1.77 \times 10^{-40}$ |
| Smk Init | Cig-per-day | 0.284 | $3.96 \times 10^{-17}$ |
| Smk Init | LfSmk | 0.869 | 0 |
| Smk Cess | Cig-per-day | 0.440 | $2.29 \times 10^{-42}$ |
| Smk Cess | LfSmk | 0.658 | $4.44 \times 10^{-157}$ |
| Cig-per-day | LfSmk | 0.545 | $1.37 \times 10^{-151}$ |
| Smoking traits vs type 2 diabetes (T2D) |  |  |  |
| Smk Init | T2D | 0.155 | $3.13 \times 10^{-19}$ |
| Smk Cess | T2D | 0.196 | $4.60 \times 10^{-12}$ |
| Cig-per-day | T2D | 0.306 | $6.96 \times 10^{-23}$ |
| LfSmk | T2D | 0.252 | $1.61 \times 10^{-43}$ |
| Smoking traits vs HbA1c |  |  |  |
| Smk Init | HbA1c | 0.067 | $1.11 \times 10^{-2}$ |
| Smk Cess | HbA1c | 0.19 | $3.98 \times 10^{-5}$ |
| Cig-per-day | HbA1c | 0.156 | $2.07 \times 10^{-5}$ |
| LfSmk | HbA1c | 0.151 | $6.30 \times 10^{-7}$ |
| Smoking traits vs body mass index (BMI) |  |  |  |
| Smk Init | BMI | 0.167 | $3.94 \times 10^{-16}$ |
| Smk Cess | BMI | 0.151 | $3.93 \times 10^{-7}$ |
| Cig-per-day | BMI | 0.243 | $2.67 \times 10^{-10}$ |
| LfSmk | BMI | 0.211 | $4.38 \times 10^{-22}$ |
| Smoking traits vs coronary artery disease (CAD) |  |  |  |
| Smk Init | CAD | 0.223 | $1.04 \times 10^{-27}$ |
| Smk Cess | CAD | 0.187 | $1.95 \times 10^{-10}$ |
| Cig-per-day | CAD | 0.294 | $1.08 \times 10^{-10}$ |
| LfSmk | CAD | 0.293 | $3.63 \times 10^{-43}$ |
| Between cardiometabolic traits |  |  |  |
| BMI | T2D | 0.491 | $4.20 \times 10^{-74}$ |
| BMI | HbA1c | 0.217 | $8.27 \times 10^{-10}$ |
| BMI | CAD | 0.232 | $2.97 \times 10^{-24}$ |
| HbA1c | T2D | 0.526 | $1.35 \times 10^{-41}$ |
| HbA1c | CAD | 0.254 | $3.15 \times 10^{-13}$ |
| T2D | CAD | 0.406 | $5.00 \times 10^{-75}$ |

**Figure 1.**
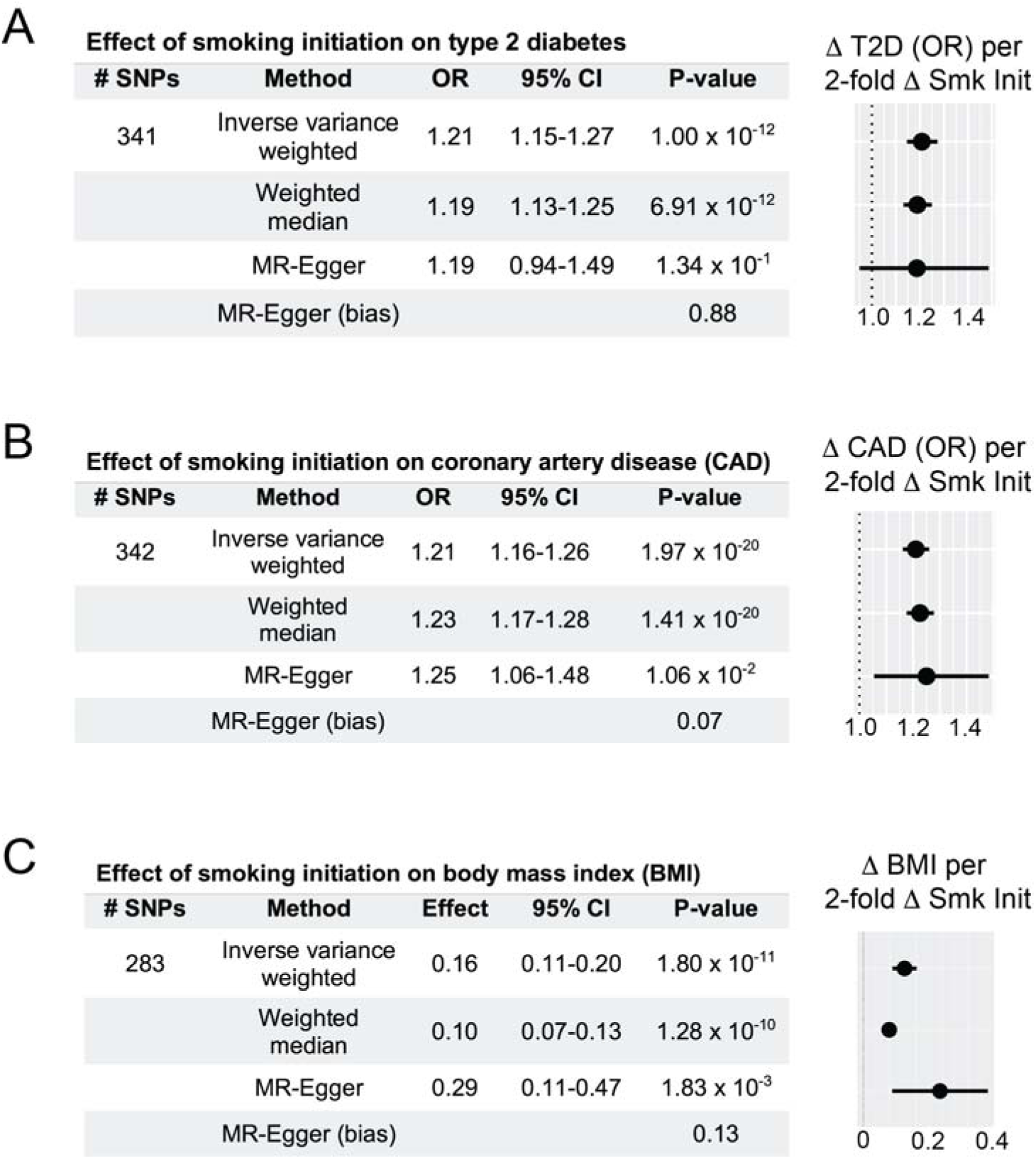
Two-sample Mendelian randomization determines that smoking initiation (Smk Init) increases type 2 diabetes (T2D) risk, coronary artery disease (CAD) risk, and body mass index (BMI). (A) Genetically determined smoking initiation risk increases T2D risk. Two-sample Mendelian randomization odds ratio (OR) estimates, 95% confidence intervals, and forest plot represent changes associated with 2-fold increase in genetic smoking initiation ‘risk’. MR-Egger intercept, a bias measurement, does not deviate significantly from zero. This validates effect estimates. (B) Increased smoking initiation risk elevates CAD risk. OR estimates, 95% confidence intervals, and forest plot represent changes associated with 2-fold increase in smoking initiation exposure. MR-Egger intercept does not deviate significantly from zero, validating the effect estimate. (**C**) Increased smoking initiation risk increases BMI. Effect estimates, 95% confidence intervals, and forest plot represent changes in BMI (standard deviation units) associated with 2-fold increase in smoking initiation exposure. MR-Egger intercept does not deviate significantly from zero, validating the effect estimate.

### Smoking cessation and heaviness were not associated with type 2 diabetes risk

We next generated instrumental variables for additional smoking behaviors, including smoking cessation and smoking heaviness (cigarettes per day). The number of instruments for both of these traits were substantially smaller (n=18 and n=42 SNPs, respectively). We did not observe evidence of association with either of these traits and type 2 diabetes risk (**Supplemental Fig. 1** and **Supplemental Table 2**). A lack of association for these smoking traits with type 2 diabetes risk may be driven by lack of statistical power to detect relatively small effect sizes. Indeed, an instrumental variable based on lifetime smoking exposure, which accounts for smoking heaviness and duration (19), identified significant effects on type 2 diabetes (**Supplemental Table 3**).

### Smoking traits are genetically correlated with type 2 diabetes risk

If the results from the above were merely due to a lack of statistical power and not a null association, we still might expect to observe positive genetic correlation between smoking behaviors and type 2 diabetes risk. Therefore, we quantified the extent to which genetic susceptibility to these smoking traits correlated with genetic susceptibility to type 2 diabetes. Using linkage disequilibrium score regression (LDSC) (20), we observed significant and positive genetic correlations between smoking traits and type 2 diabetes (**Table 1**). These results suggest that future genetic studies that explain more of the genetic variance in smoking behavior and type 2 diabetes might clarify the effects of these smoking traits on type 2 diabetes.

### Smoking initiation may increase HbA1c levels

As a sensitivity analysis, we next estimated the causal effect of smoking initiation on genetically determined HbA1c levels (21). HbA1c level represents glycated hemoglobin and is used to clinically diagnose type 2 diabetes. As such, HbA1c level is a biochemical surrogate for type 2 diabetes. We created a genetic instrument comprised of 282 SNPs, and observed that a 2-fold increase in genetically determined smoking risk was associated with a 0.02 standard deviation unit increase in HbA1c level (95% CI: 0.01-0.03, P=3.1×10^−3^ by inverse variance weighted method, **Supplemental Fig. 2**). Although this association was not significant in weighted median or MR-Egger tests, these findings gave some additional support for a relationship between smoking behavior and glycemic regulation (4,8), and between genetically determined smoking risk and type 2 diabetes.

**Figure 2.**
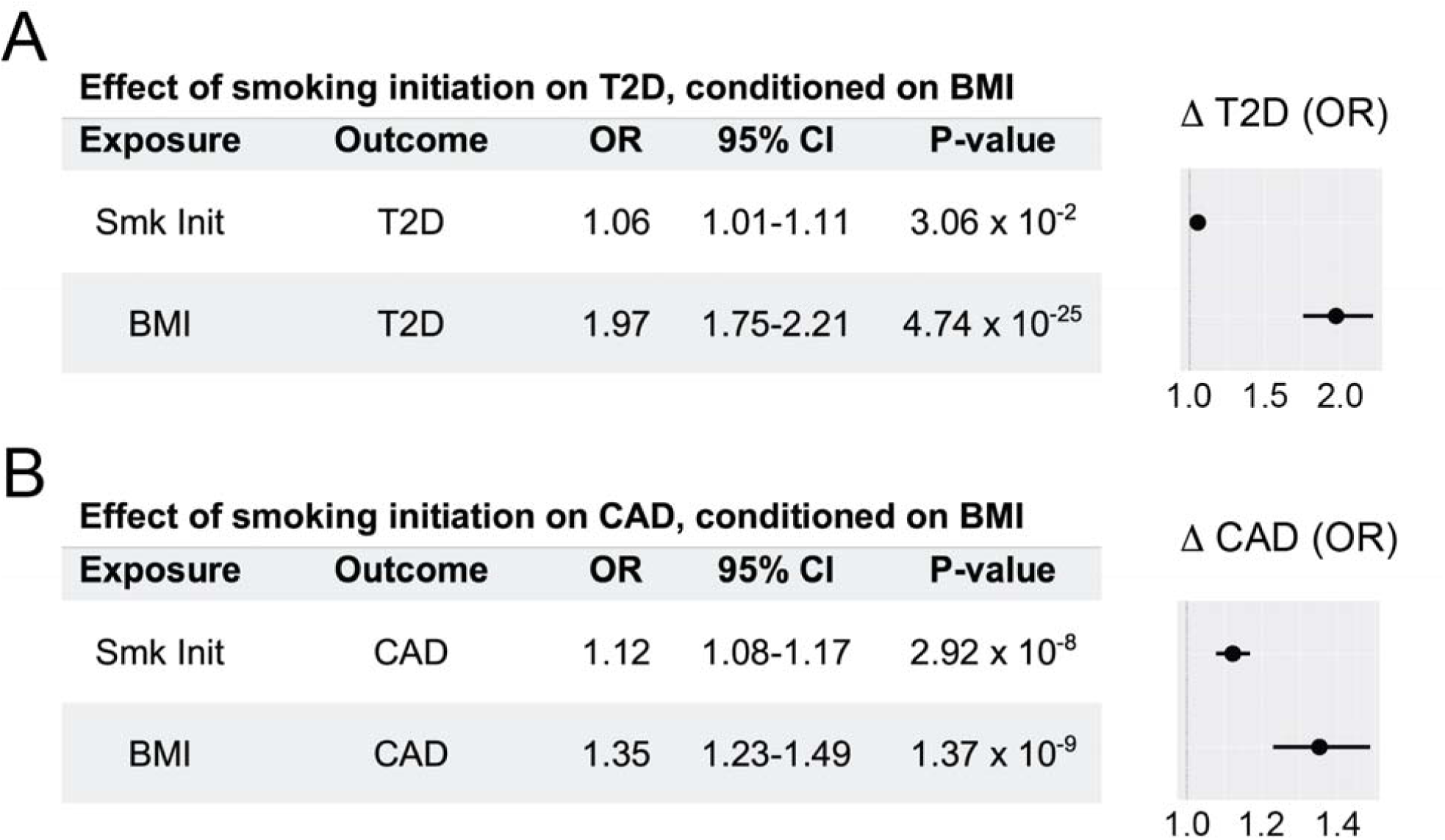
Body mass index (BMI) largely mediates the effect of smoking initiation on increased type 2 diabetes (T2D) risk, but not coronary artery disease (CAD) risk. Instrumental variables for these experiments comprised ∼286 SNPs from smoking initiation GWAS summary statistics (16).(**A**) Multivariable mendelian randomization (MVMR) results show that genetically determined BMI largely confounds the effect of smoking initiation on T2D risk. (**B**) Multivariable mendelian randomization (MVMR) results show that smoking initiation retains a strong independent effect on CAD risk after conditioning on BMI. Odds ratio (OR) estimates, 95% confidence intervals, and forest plots represent changes in outcomes associated with 2-fold increase in genetic smoking initiation ‘risk’, conditioned on BMI.

### Increased risk of type 2 diabetes by smoking initiation is equivalent to that of coronary artery disease

To contrast our estimated effect with established causal relationships, we next used MR to address the effect of smoking initiation on susceptibility to coronary artery disease (CAD). We observed a positive causal effect from smoking on CAD risk (**Fig. 1B** and **Supplemental Table 3**), with a 2-fold increase in smoking initiation risk increasing odds of CAD risk by 21% (95% CI: 1.16-1.26, P=2.0×10^−20^). This estimated causal effect on CAD was consistent with previous reports (5). Strikingly, the estimated effect of smoking initiation on CAD risk was nearly identical to increased type 2 diabetes risk from this behavior (**Fig. 1A**). These results suggested that smoking initiation portends an equivalently increased risk of type 2 diabetes and CAD. However, it remained to be seen whether biological factors shared between diseases mediate the observed associations.

### Genetically elevated smoking initiation risk is associated with increased body mass index

Prior evidence has demonstrated a positive correlation between smoking and body mass index (BMI) (9,11), although some genetic results based on restricted set of loci indicated that BMI might actually decrease as a result of smoking (10). This latter finding is consistent with reported clinical observations that linked smoking cessation with weight gain (22). We hypothesized that BMI mediates the effects of smoking on type 2 diabetes and/or CAD risk.

We first tested the causal association of BMI with smoking initiation and demonstrated that a 1,364 SNP instrumental variable based on BMI (23) increased smoking initiation risk, with a 25% increase in odds of smoking initiation per standard deviation unit increase in genetically determined BMI (95% CI: 1.21-1.29, P=2.1×10^−44^, **Supplemental Fig. 3**). We noted that the MR-Egger intercept significantly deviated from zero, suggesting the presence of directional pleiotropy (P=1.1×10^−4^, **Supplemental Fig. 3**). Results using methods more robust to the presence of pleiotropy (e.g., weighted median and MR-Egger) demonstrated significant effects of smoking on BMI, though effect estimates differed (**Supplemental Fig. 3**). Overall, these results confirmed the previously reported genetic association (9,11).

**Figure 3.**
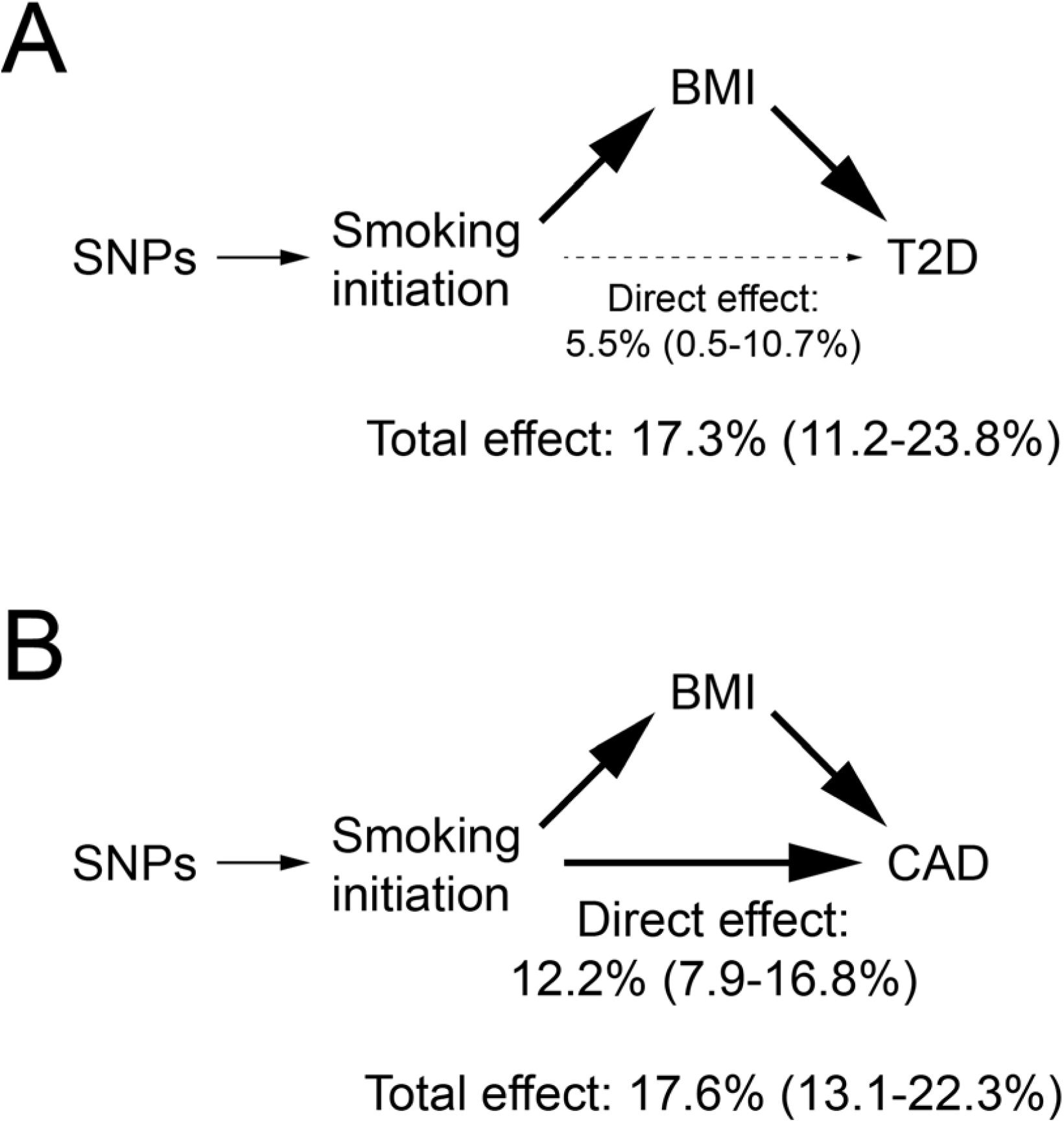
Models for how genetically determined smoking initiation risk impacts cardiovascular disease traits. (**A**) Single nucleotide polymorphisms (SNPs) determine genetic risk of smoking initiation. By MR-Steiger estimates (25), smoking initiation directionally influences BMI. From mediation analysis (15), the total and direct effects of smoking initiation on type 2 diabetes risk (T2D) are shown, representing increased odds of T2D (with 95% confidence interval) per 2-fold increase in genetically determined smoking initiation risk. (**B**) Both smoking initiation and BMI have strong independent effects on CAD risk. Total and direct effects of smoking initiation on coronary artery disease risk (CAD) are shown, representing increased odds of CAD (with 95% confidence interval) per 2-fold increase in genetically determined smoking initiation risk.

Next, we conversely assessed whether genetic risk of initiating smoking behavior was associated with BMI. Indeed, there was a positive association between smoking and BMI (**Fig. 1C** and **Supplemental Table 3**), with each 2-fold increase in genetically determined smoking initiation risk corresponding to 0.16 standard deviation unit increase in BMI (95% CI: 0.11-0.20, P=1.8×10^−11^). We noted that the MR-Egger intercept was not significantly different from zero (P=0.13), indicating little if any evidence of directional bias on this estimated effect (**Fig. 1C**). Our findings indicated a significant, shared genetic risk between smoking and BMI. From a biological standpoint, these results supported prior studies relating nicotine craving to a desire to overeat (11).

### BMI mediates the effect of smoking initiation risk on type 2 diabetes

We reasoned that smoking effects on BMI could mediate the potentially complex relationship between smoking and type 2 diabetes. We therefore estimated causal effects of smoking on type 2 diabetes risk, accounting for BMI, using multivariable MR. Our results showed that the effects of smoking on type 2 diabetes are largely explained by BMI. When conditioned on BMI, the residual independent (direct) effect of smoking on type 2 diabetes was attenuated, with an estimated OR 1.06 (95% CI: 1.01-1.11, P=0.03, **Fig. 2A**). In an analogous experiment, we found that the effect of smoking initiation on HbA1c was no longer significant (P= 0.24, **Supplemental Fig. 4**). Further, an instrument based on lifetime smoking exposure, accounting for smoking heaviness and duration (19), showed qualitatively similar results (OR 1.25, CI: 0.97-1.62, P=0.09, **Supplemental Fig. 5A**).

We next considered the reciprocal experiment, using an instrumental variable (>1,300 SNPs) based on BMI, to estimate effects of BMI whilst accounting for smoking behavior on type 2 diabetes susceptibility or HbA1c levels (**Supplemental Fig. 6A-B**). BMI is an established, causal risk factor for type 2 diabetes (24), and the traits share a common genetic basis (**Table 1**). As expected, we found that a 1 standard deviation increase in genetically determined BMI corresponded to a 2.5-fold increase in diabetes risk (95% CI: 2.36-2.70, P=6.6×10^−134^).

However, smoking was not associated with T2D (P=0.54). Similarly, a 1 standard deviation increase in genetically determined BMI was associated with a 0.06 unit increase in HbA1c (95% CI: 0.05-0.07, P=4.2×10^−24^) but smoking initiation was not associated with HbA1c levels (P=0.14). These results demonstrate that BMI accounts for most if not all of the effect of genetically determined smoking propensity on increased type 2 diabetes risk.

### BMI does not mediate the effect of smoking initiation risk on CAD

We then asked whether the effects of smoking on CAD were similarly mediated by BMI. As with BMI and type 2 diabetes, we observed significant genetic correlation between BMI and CAD (r_g_=0.23, P=3.0×10^−24^, **Table 1**). In contrast to type 2 diabetes, there remained a significant independent effect of smoking initiation on CAD risk after controlling for BMI (OR=1.12, 95% CI: 1.08-1.17, P=2.9×10^−8^, **Fig. 2B**). Analyses using genetic instruments based on lifetime smoking exposure (OR=1.72, CI: 1.43-2.07, P=9.2×10^−8^, **Supplemental Fig. 5B**) and BMI (OR=1.23, CI: 1.14-1.32, P=7.9×10^−8^, **Supplemental Fig. 6C**) both showed a persistent effect of smoking on susceptibility to CAD. This suggests that the biology underlying the association between smoking and CAD partially involves obesity-related pathways, but is largely due to direct effects of smoking on atherosclerosis and/or contributions from other mediating factors.

### Smoking increases type 2 diabetes risk via BMI, and independently increases CAD risk

We sought to assess the directionality of the estimated causal effect between smoking, BMI and type 2 diabetes. MR-Steiger can infer the direction of causality using GWAS summary statistics in situations where the biology of underlying SNPs are not yet understood (25). We used MR-Steiger to test whether the multi-trait relationships seen in our analyses were best explained by smoking initiation or BMI as driving associations with type 2 diabetes. For this experiment, we analyzed 1085 SNPs that met genome-wide significance (P<5×10^−8^) in GWAS for smoking initiation and BMI. We determined that our findings were best explained by a model in which smoking increased BMI (MR-Steiger sensitivity 14.95, P<10^−10^, **Fig. 3**).

Finally, we used mediation analyses to estimate the total and direct effects of smoking on type 2 diabetes and CAD risks (15). In mediation analysis, binary outcomes preclude accurate estimation of indirect effects (15). We found that the total effect of a 2-fold genetically increased smoking initiation risk was to increase the odds of type 2 diabetes risk by 17.3% (95% CI: 11.2-23.8%), although the direct effect attributable to smoking initiation was just 5.5% (95% CI: 0.5-10.7%, **Fig. 3A**). In contrast, the total effect of increased smoking initiation was to increase CAD risk by 17.6% (CI: 13.1-22.3%), with a direct effect from smoking initiation of 12.2% (95% CI: 7.9-16.8%, **Fig. 3B**). Mediation analyses based on lifetime smoking score were consistent with these effect patterns (**Supplemental Table 4**). In sum, these results demonstrated that the majority of the effect from smoking on type 2 diabetes was mediated by BMI, whereas both smoking behavior and BMI independently affect CAD risk.

## Discussion

Despite long-standing clinical observations (5), a causal genetic association between smoking and type 2 diabetes has only recently been established (7). Our results indicate a more complex relationship between smoking and type 2 diabetes than what has been reported previously (1,6,7,26). We associated a 2-fold increased smoking initiation risk with a BMI increase of 0.16 standard deviation units, or ∼0.8 kg/m^2^ (27). Genetically determined smoking initiation risk increases type 2 diabetes risk and HbA1c, but these associations are mostly mediated through an association between smoking on BMI. Our findings were consistent across instrumental variable methodological approaches, and we did not observe evidence of systematic biases to our effect estimate. By clarifying the biological factors underlying these trait associations, our findings will allow for more informed clinical recommendations and improve targeted therapeutic development.

Our findings add to numerous reports regarding the negative effects of smoking on cardiometabolic health identified by medical practitioners and policymakers. A recent report detailed one mechanism directly linking smoking with increased blood glucose and type 2 diabetes risk (28). We anticipate that important biological pathways for this association will involve altered obesity and/or adiposity. Indeed, our findings indicate that future validation research should focus on such pathways to establish relevant biological mechanisms.

The association between smoking and type 2 diabetes was similar in magnitude to the association between smoking and coronary artery disease (CAD). Interestingly, the effect of smoking on CAD risk were not mediated by BMI. Although outside the scope of our current study, it will be important for future work to identify how genetically determined smoking risk portends increased CAD risk. Indeed, smoking has myriad deleterious effects, including but not limited to blood pressure elevation, endothelial damage, and enhanced inflammation (29). Direct effects from smoking on atherosclerosis may underlie this strong genetic association. Further revealing alternative pathways and risk factors that account for these findings may have translational benefit for diagnosing and treating heart disease in smokers.

In sum, our findings reinforce the importance of clinical recommendations to avoid or stop tobacco smoking. By elucidating genetic factors mediating the associations between smoking behavior, BMI, type 2 diabetes and CAD, our results will inform strategies to mitigate these risks though development of novel therapeutics. Although interpretation of these findings is limited to individuals of European ancestry, we expect that well-powered multi-ethnic GWAS will ultimately confirm this result in other populations.

## Materials and Methods

### Genome-wide summary statistics collection

We analyzed publicly available GWAS summary statistics for smoking initiation (n=1.2 million individuals), smoking cessation (n=547,219), cigarettes per day (n=337,334) (16), type 2 diabetes (n=898,130) (17), HbA1c (n=123,665) (21), body mass index (BMI, n=∼700,000) (23), and coronary artery disease (CAD, n=∼546,000) (30). Lifetime smoking index was based on UK Biobank data (n=462,690) (19). Despite some sample overlap across these GWAS, the risk of sample overlap bias is low given the large size of these major international consortia (31). We used data from individuals of European ancestry only. All data sets were analyzed in genome build hg19.

Smoking initiation was defined as having smoked ‘regularly’, every day for at least 1 month, or >100 cigarettes in one’s lifetime (16). Smoking cessation was defined as those who were identified as having initiated smoking but subsequently stopped. Smoking heaviness (cigarettes per day) was defined as the number of cigarettes smoked per day, on average, as a current or former smoker. Lifetime smoking index scores, which take into account smoking initiation, heaviness and duration, were previously calculated (19).

### Genetic variant selection related to smoking exposures

We created genetic instrumental variables (IVs) for smoking traits, including ‘smoking initiation’, ‘smoking cessation’, ‘cigarettes per day’ (smoking heaviness) (16), and lifetime smoking exposure (19), as well as HbA1c (21) and BMI (23). To generate IVs, we first identified SNPs common to both exposure and outcome data sets. Using Two-sample MR, we then clumped all genome-wide significant SNPs to identify single nucleotide polymorphisms within independent linkage disequilibrium blocks (EUR r^2^<0.01) in 250kb regions. Full data sets for our IVs are shown in **Supplemental Tables 5-22**.

We used mRnd (http://cnsgenomics.com/shiny/mRnd/, (18)) to estimate the strength F-statistics of these IVs. Smoking initiation or cessation were input as binary exposure variables. All other traits were analyzed as continuous exposure variables. We calculated proportion of genetic inheritance explained in each exposure per Shim *et. al*. (32). None of our instrumental variables was subject to weak instrument bias, as each had an F-statistic greater than 10 (**Supplemental Table 1**).

### Mendelian randomization and causal effect estimation

We performed two-sample MR (TwoSample MR package v0.5.2 (13)) using R (v3.6.1). We present causal estimates from inverse variance weighted (random effects model), weighted median, and MR-Egger regression methods. We analyzed for pleiotropic bias using MR-Egger regression intercepts, wherein significant non-zero intercepts can imply directional bias among IVs (33). We performed multivariable Mendelian randomization analyses using the MVMR package (14) in R, and present causal estimates for each associated variable. For causal direction analysis, we used MR-Steiger and report values for sensitivity, statistical significance, and ‘correct causal direction’ (25).

### Genetic correlation estimates

Genetic correlations were estimated using Linkage Disequilibrium Score Regression (LDSC) (20). Summary statistics were munged and analyzed for each trait. Presented data reflecting genetic correlation estimates (i.e., r_g_ values) and related statistical significance estimates (p-values).

### Statistical analysis

In MR analyses, estimated effects from exposure(s) on outcome are presented from inverse variance weighted, weighted median, and MR-Egger regression measures. Because Cochran’s Q test (included in the TwoSample MR package v0.5.2 (13)) found heterogeneity our IVs, we utilized the random-effect model when performing inverse-variance weighted MR. We also performed a sensitivity analysis using IV pruning via MR-PRESSO (34) for a single trait pair (smoking initiation and type 2 diabetes). The resultant IV included 325 SNPs from an initial 341, but MR results were not qualitatively different than the full instrument (data not shown). Thus, we performed and report MR results using IVs that had not undergone pruning. Statistical significance was defined as P < 0.05 for all experiments.

For continuous outcomes (BMI and HbA1c), results are presented as beta effect values representing changes in standard deviation units for these traits. For the dichotomous outcomes smoking initiation, type 2 diabetes, and CAD, we converted causal effect estimates into odds ratios using previously described methods (35). By multiplying causal effect estimates by ln(2) and taking the exponentiated value (=exp^[ln(2)*effect]), we calculated the change in outcome odds ratio that corresponded with a 2-fold change in genetically determined dichotomous exposure risk. For continuous exposure variables, we exponentiated the causal effect estimate (=exp^[effect]) to reach a value reflecting the change in outcome per standard deviation unit increase in exposure.

### Mediation analysis

Mediation analysis estimates were calculated as described by Burgess *et al* (15). The outcome variables analyzed (type 2 diabetes and CAD risk) are defined in non-collapsible odds ratios.

We therefore present only total and direct effects. Indirect effects are inaccurate since there cannot be linear relationships between exposures, mediators, and these binary outcomes (15).

### Coding scripts and data sets

All relevant coding scripts and data sets can be found on Github (https://github.com/thomchr/SmkT2D). All data and coding scripts are also available upon request.

## Data Availability

All data used for this work utilize summary data that is available in the public domain.

## Acknowledgements

CST and BFV designed the study. All authors conducted, analyzed, and/or interpreted experimental data. CST and BFV wrote the paper. BFV acts as guarantor and corresponding author for this manuscript.

This work was supported by National Institutes of Health [DK101478 and HG010067 to BFV, HD043021 to CST], an American Academy of Pediatrics Marshall Klaus Neonatal-Perinatal Research Award to CST, and a Linda Pechenik Montague Investigator Award to BFV. SMD is supported by the US Department of Veterans Affairs, Clinical Science Research and Development (IK2-CX001780). This publication does not represent the views of the Department of Veterans Affairs or the US Government. All authors confirm independence from funders and that all authors had full access to all of the statistics in the study. As such they can attest to the integrity of the data and the accuracy of the data analysis.

## Supplemental Material

### Supplemental Figures

**Supplementary Figure 1.**
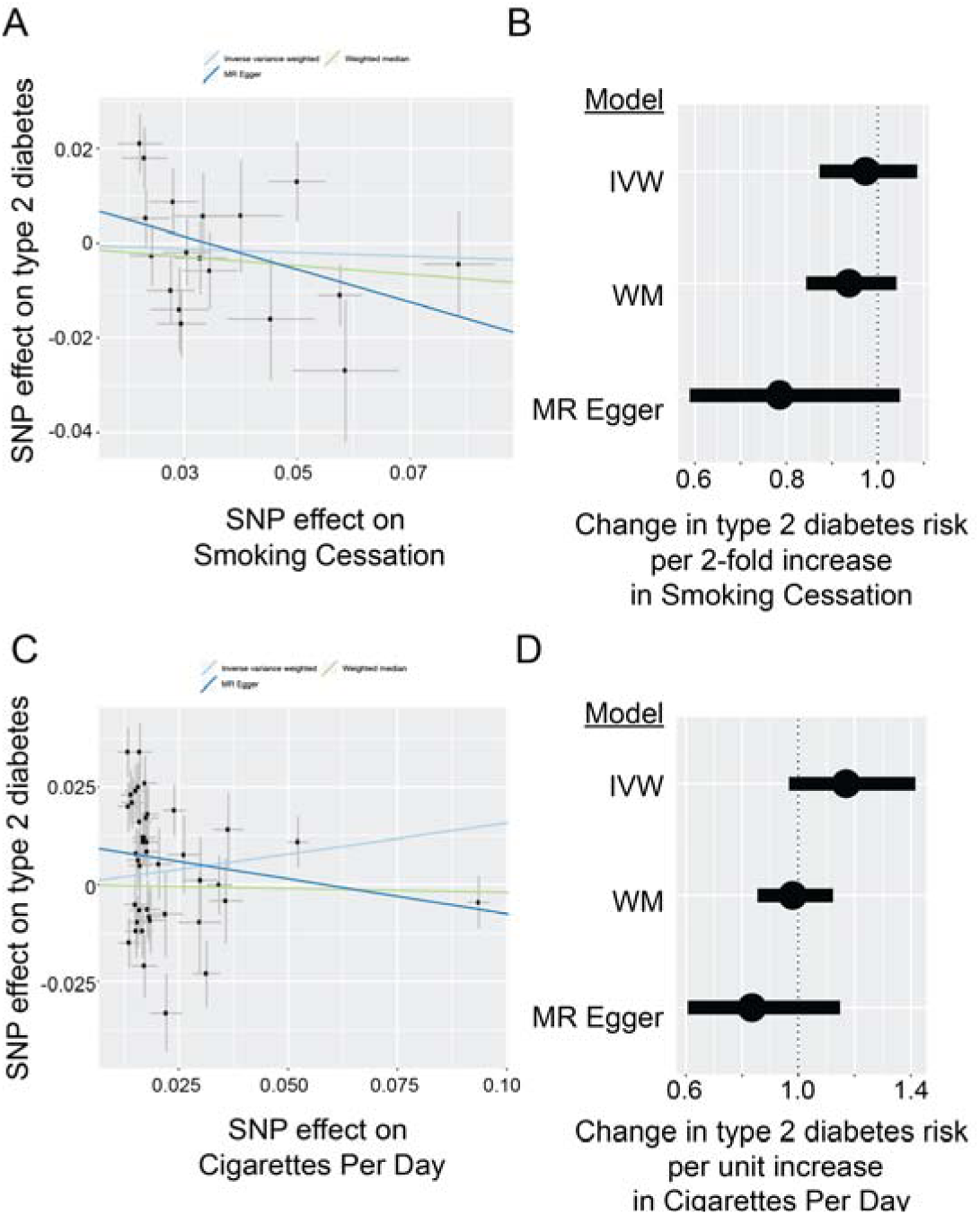
Two-sample mendelian randomization shows no significant effects of smoking cessation or smoking heaviness on type 2 diabetes risk. (**A**) Using an 18-SNP instrumental variable for smoking cessation, two-sample MR does not define an increased risk of type 2 diabetes. Scatter plot shows effect sizes (mean ± standard error) for smoking cessation and type 2 diabetes for each SNP in the instrumental variable, along with line of best fit for inverse variance weighted, weighted median, and MR-Egger regression models. (**B**) Forest plot showing aggregated effects for smoking cessation on type 2 diabetes, shown as change in type 2 diabetes risk (odds ratio) per 2-fold change in smoking cessation “risk” (mean ± standard error) for inverse variance weighted (IVW), weighted median (WM), and MR-Egger regression models. (**C**) Using a 42-SNP instrumental variable for smoking heaviness (number of cigarettes per day), two-sample MR does not define an increased risk of type 2 diabetes. Scatter plot shows effect sizes (mean ± standard error) for smoking heaviness and type 2 diabetes for each SNP in the instrumental variable, along with line of best fit for inverse variance weighted, weighted median, and MR-Egger regression models. (**D**) Forest plot showing aggregated instrumental variable effects for smoking heaviness on type 2 diabetes, shown as change in type 2 diabetes risk (odds ratio) per unit increase in smoking heaviness (mean ± standard error) for inverse variance weighted (IVW), weighted median (WM), and MR-Egger regression models.

**Supplementary Figure 2.**
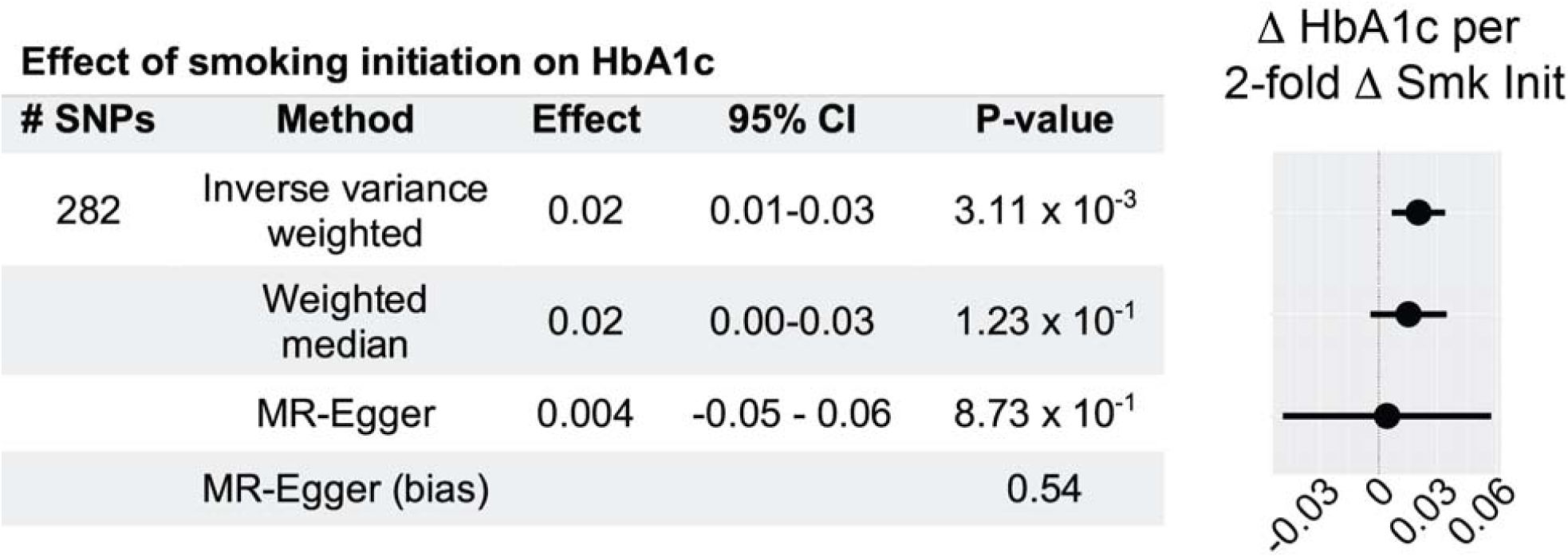
Increased smoking initiation risk elevates HbA1c by inverse variance weighted method. Effect estimates, 95% confidence interval, and forest plot represent changes in HbA1c (standard deviation units) associated with 2-fold increase in smoking initiation exposure. MR-Egger intercept does not deviate significantly from zero, validating the effect estimate.

**Supplementary Figure 3.**
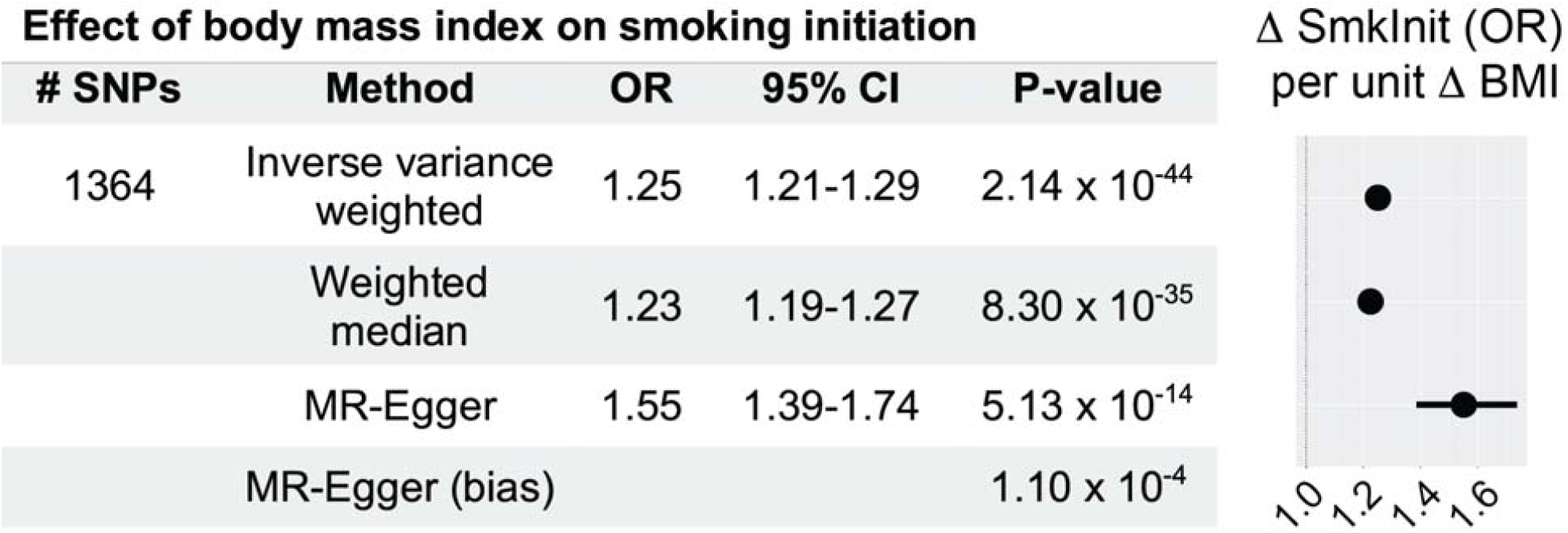
Genetically determined body mass index (BMI) is associated with increased smoking initiation. Effect estimates, 95% confidence intervals, and forest plot for smoking initiation ‘risk’ represent changes in odds ratio (OR) for smoking initiation per unit increase in BMI. The MR-Egger intercept deviates significantly from zero, invalidating the effect estimate. The statistically significant effects seen remain valid.

**Supplementary Figure 4.**
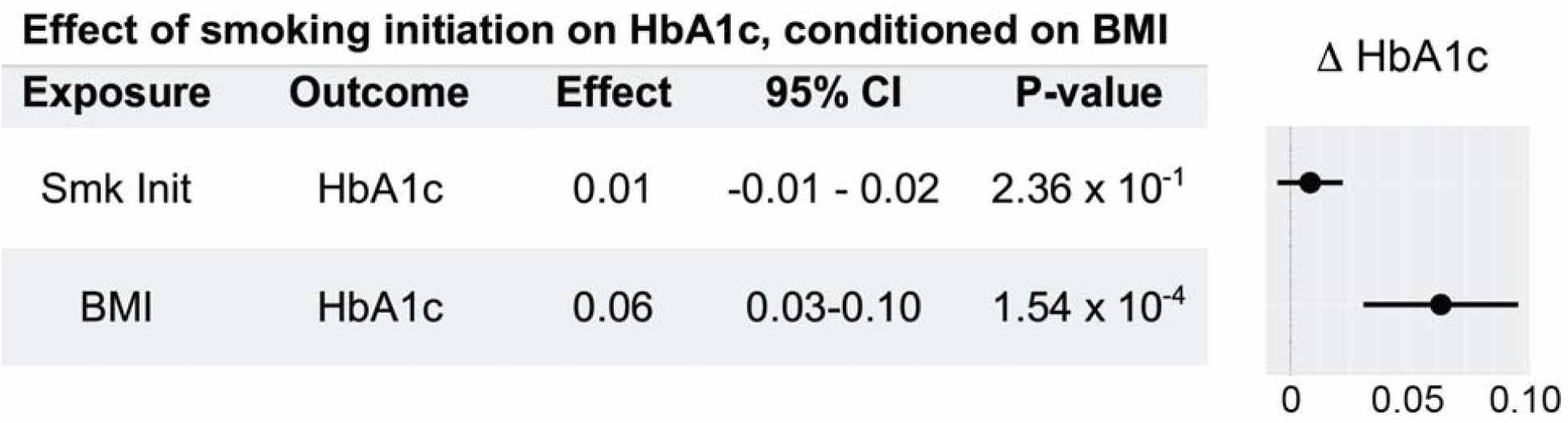
Body mass index (BMI) mediates the effect of smoking initiation on HbA1c. Multivariable mendelian randomization (MVMR) results show that genetically determined BMI accounts for the effect of smoking initiation on HbA1c. Effect estimates, 95% confidence intervals, and forest plot represent changes in HbA1c (standard deviation units) per 2-fold increase in genetically determined smoking initiation risk, conditioned on BMI.

**Supplementary Figure 5.**
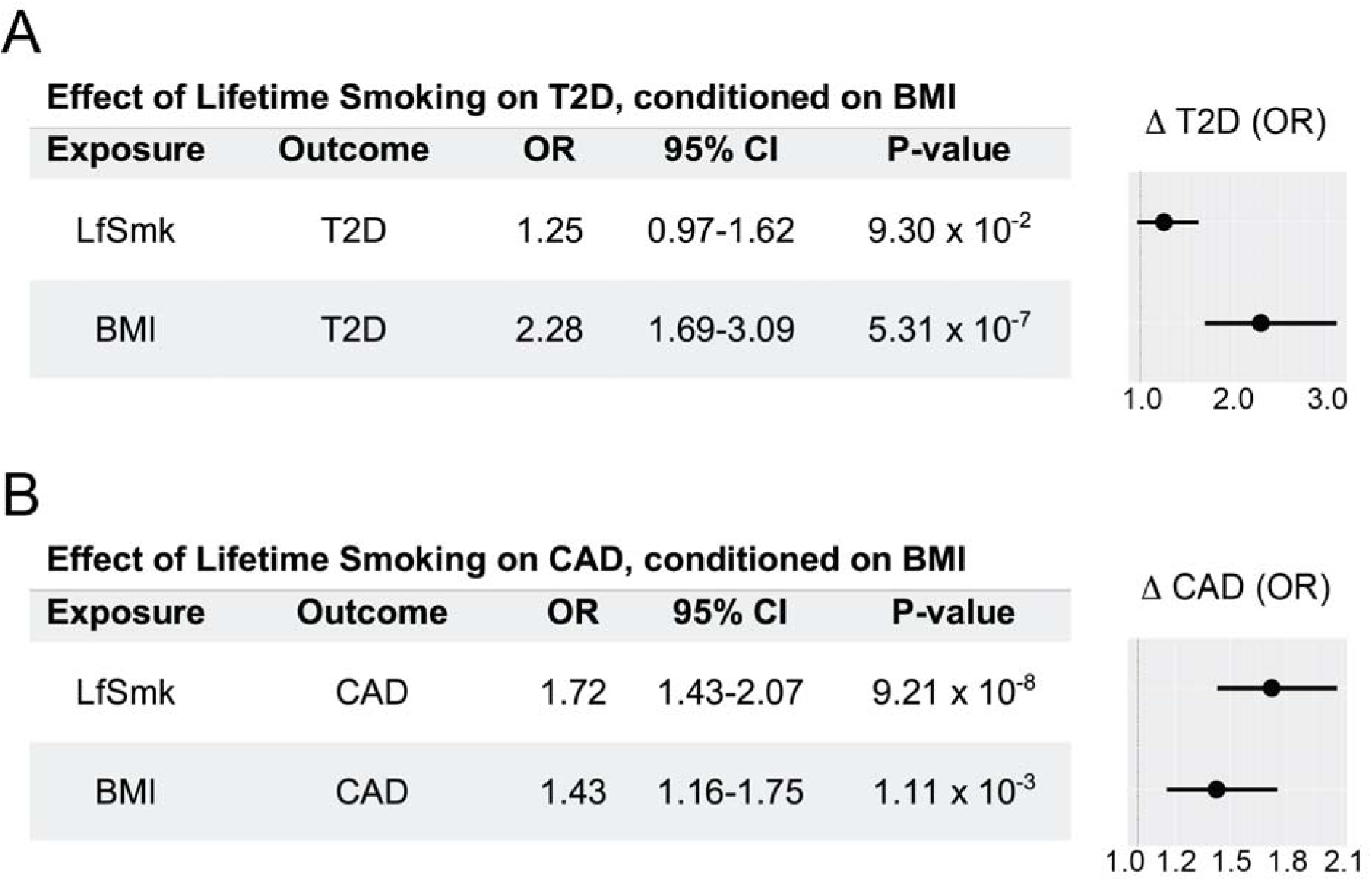
Body mass index (BMI) mediates the effect of smoking, as defined by Lifetime Smoking score (LfSmk), on increased type 2 diabetes (T2D) risk, but not coronary artery disease (CAD) risk. Instrumental variables for these experiments comprised 101 SNPs with calculated Lifetime smoking scores (19). (**A**) Multivariable mendelian randomization (MVMR) results show that genetically determined BMI accounts for the effect of lifetime smoking score (LfSmk) on T2D risk. (**B**) Multivariable mendelian randomization (MVMR) results show that BMI and LfSmk have independent effects on CAD risk. Effect and odds ratio (OR) estimates, 95% confidence intervals, and forest plots represent changes per standard deviation unit increase in LfSmk, conditioned on BMI.

**Supplementary Figure 6.**
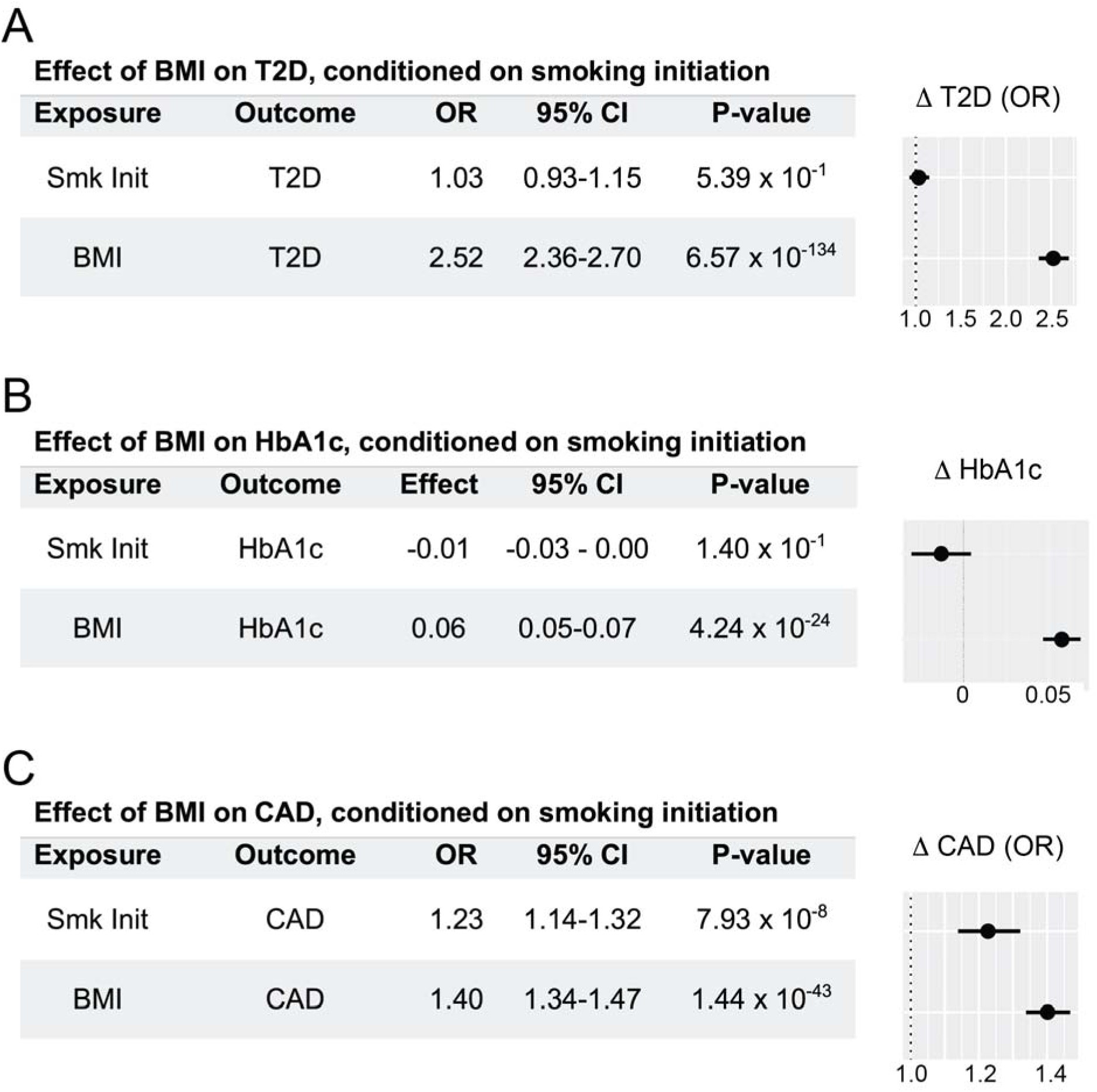
Body mass index (BMI) mediates the effect of smoking initiation on increased type 2 diabetes (T2D) risk, but not coronary artery disease (CAD) risk. Instrumental variables for these experiments comprised ∼1385 SNPs from BMI GWAS summary statistics (23). (**A**) Multivariable mendelian randomization (MVMR) results show that genetically determined BMI accounts for the effect of smoking initiation on T2D risk. (**B**) Multivariable mendelian randomization (MVMR) results show that genetically determined BMI accounts for the effect of smoking initiation on HbA1c (standard deviation units). (**C**) Multivariable mendelian randomization (MVMR) results show that BMI and smoking initiation have independent effects on CAD risk. Effect and odds ratio (OR) estimates, 95% confidence intervals, and forest plots represent changes per unit increase in genetically determined BMI, conditioned on smoking initiation risk.

### Supplemental Tables Legends

**Supplemental Table 1**.

Instrument strength F-statistics and related values from GWAS necessary to perform calculations for Two-sample Mendelian randomization experiments (http://cnsgenomics.com/shiny/mRnd/, (18)). We calculated genetic risk explained in each exposure using a previously described method Shim *et al* (32).

**Supplemental Table 2**.

Smoking cessation and smoking heaviness (cigarettes per day, Cig per day) do not reach statistically significant effects on type 2 diabetes risk based on inverse variance weighted (IVW), weighted median (WM) or MR-Egger regression metrics. Odds ratio (OR) effects and 95% confidence intervals represent change in type 2 diabetes OR associated with 2-fold increased smoking cessation risk, or change in type 2 diabetes OR per unit increase in smoking heaviness (Cig per day). Significant MR-Egger regression intercept values that deviate significantly from zero (e.g., Cig per day) invalidate effect estimates.

**Supplemental Table 3**.

Two-sample Mendelian randomization analysis results for Lifetime smoking score (LfSmk (19)) on type 2 diabetes, coronary artery disease, and body mass index. Odds ratio (OR)/Effect and 95% confidence intervals represent changes in disease risk or body mass index associated with a 1 standard deviation unit increase in LfSmk score. Significant MR-Egger regression intercept values that deviate significantly from zero (e.g., type 2 diabetes) invalidate effect estimates.

**Supplemental Table 4**.

Mediation analyses depicting total and direct effects of lifetime smoking score (LfSmk) on type 2 diabetes (T2D) or coronary artery disease (CAD) (15). Values represent increased disease risk (odds ratio, OR) per 1 standard deviation increase in LfSmk score.

**Supplemental Table 5**.

Instrumental variable data for MR experiments estimating effects of smoking initiation on type 2 diabetes. The rsid (hg19), chromosome, position, effect allele, non-effect allele, effect sizes and standard errors are shown for each SNP.

**Supplemental Table 6**.

Instrumental variable data for MR experiments estimating effects of smoking cessation on type 2 diabetes. The rsid (hg19), chromosome, position, effect allele, non-effect allele, effect sizes and standard errors are shown for each SNP.

**Supplemental Table 7**.

Instrumental variable data for MR experiments estimating effects of smoking heaviness (cigarettes per day) on type 2 diabetes. The rsid (hg19), chromosome, position, effect allele, non-effect allele, effect sizes and standard errors are shown for each SNP.

**Supplemental Table 8**.

Instrumental variable data for MR experiments estimating effects of smoking initiation on HbA1c. The rsid (hg19), chromosome, position, effect allele, non-effect allele, effect sizes and standard errors are shown for each SNP.

**Supplemental Table 9**.

Instrumental variable data for MR experiments estimating effects of smoking initiation on coronary artery disease (CAD). The rsid (hg19), chromosome, position, effect allele, non-effect allele, effect sizes and standard errors are shown for each SNP.

**Supplemental Table 10**.

Instrumental variable data for MR experiments estimating effects of smoking initiation on BMI. The rsid (hg19), chromosome, position, effect allele, non-effect allele, effect sizes and standard errors are shown for each SNP.

**Supplemental Table 11**.

Instrumental variable data for MR experiments estimating effects of BMI on smoking initiation. The rsid (hg19), chromosome, position, effect allele, non-effect allele, effect sizes and standard errors are shown for each SNP.

**Supplemental Table 12**.

Instrumental variable data for MR experiments estimating effects of lifetime smoking score (LfSmk) on type 2 diabetes (T2D). The rsid (hg19), chromosome, position, effect allele, non-effect allele, effect sizes and standard errors are shown for each SNP.

**Supplemental Table 13**.

Instrumental variable data for MR experiments estimating effects of lifetime smoking score (LfSmk) on body mass index (BMI). The rsid (hg19), chromosome, position, effect allele, non-effect allele, effect sizes and standard errors are shown for each SNP.

**Supplemental Table 14**.

Instrumental variable data for MR experiments estimating effects of lifetime smoking score (LfSmk) on coronary artery disease (CAD). The rsid (hg19), chromosome, position, effect allele, non-effect allele, effect sizes and standard errors are shown for each SNP.

**Supplemental Table 15**.

Instrumental variable data for MR experiments estimating effects of lifetime smoking on T2D, conditioned on BMI. The rsid (hg19), chromosome, position, effect allele, non-effect allele, effect sizes and standard errors are shown for each SNP.

**Supplemental Table 16**.

Instrumental variable data for MR experiments estimating effects of lifetime smoking on CAD, conditioned on BMI. The rsid (hg19), chromosome, position, effect allele, non-effect allele, effect sizes and standard errors are shown for each SNP.

**Supplemental Table 17**.

Instrumental variable data for MR experiments estimating effects of smoking initiation on HbA1c, conditioned on BMI. The rsid (hg19), chromosome, position, effect allele, non-effect allele, effect sizes and standard errors are shown for each SNP.

**Supplemental Table 18**.

Instrumental variable data for MR experiments estimating effects of smoking initiation on T2D, conditioned on BMI. The rsid (hg19), chromosome, position, effect allele, non-effect allele, effect sizes and standard errors are shown for each SNP.

**Supplemental Table 19**.

Instrumental variable data for MR experiments estimating effects of smoking initiation on CAD, conditioned on BMI. The rsid (hg19), chromosome, position, effect allele, non-effect allele, effect sizes and standard errors are shown for each SNP.

**Supplemental Table 20**.

Instrumental variable data for MR experiments estimating effects of BMI on T2D, conditioned on smoking initiation. The rsid (hg19), chromosome, position, effect allele, non-effect allele, effect sizes and standard errors are shown for each SNP.

**Supplemental Table 21**.

Instrumental variable data for MR experiments estimating effects of BMI on HbA1c, conditioned on smoking initiation. The rsid (hg19), chromosome, position, effect allele, non-effect allele, effect sizes and standard errors are shown for each SNP.

**Supplemental Table 22**.

Instrumental variable data for MR experiments estimating effects of BMI on CAD, conditioned on smoking initiation. The rsid (hg19), chromosome, position, effect allele, non-effect allele, effect sizes and standard errors are shown for each SNP.

